# Cardiac tissue perfusion at rest correlates with systemic microvascular function in those with and without atherosclerotic coronary artery disease; a cross-sectional study

**DOI:** 10.1101/2024.12.10.24318777

**Authors:** Joshua Wilcox, Oliver Gosling, Francesco Casanova, Jingzhou He, Kim M Gooding, Andrew Pitt, Claire Ball, Gareth Morgan-Hughes, Nick Bellenger, Angela C Shore, W David Strain

**Author notes:** Corresponding Author*: Prof David Strain Diabetes and Vascular Medicine, University of Exeter, College of Medicine and Health, Royal Devon & Exeter Hospital, Barrack Road, Exeter, EX2 5AX. Joint first authors (Joshua Wilcox, Oliver Gosling).

## Abstract

**Background:** Coronary microvascular disease is often defined by symptoms in the absence of epicardial coronary artery stenosis. There is, however, a growing interest in exploring the vascular physiology of patients with chest pain syndromes who have been confirmed to have unobstructed coronary arteries. As it is known that people with microvascular coronary disease have an additive poor prognosis, we aimed to determine whether this was part of a systemic microvascular dysregulation. As such, we explored the correlations between cardiovascular magnetic resonance (CMR) myocardial perfusion with cutaneous maximal hyperaemic response (MHR) and post-occlusive reactive hyperaemia (PORH), as assessed by laser Doppler fluximetry, in patients with known coronary anatomy determined via computed tomography coronary angiography (CTCA).

**Methods:** MHR was measured in response to local heating to 42°C and PORH was measured in response to a 4-minute ischaemic stimulus in 102 participants with and without diabetes and/or coronary artery disease, defined as coronary artery calcification of >0 Agatston units. Subepicardial and subendocardial perfusion at rest and in response to adenosine stress was measured via CMR.

**Results:** Out of 102 participants, 47 (45.1%) had diabetes, and 59 (57.8%) had coronary artery disease, with 32 (31.4%) having both. MHR and PORH were attenuated in participants with diabetes. Resting, but not stressed, CMR perfusion in all subepicardial and subendocardial territories was proportionately impaired in those with attenuated MHR. This association was independent of conventional risk factors including age, sex, blood pressure, glycaemia, coronary artery disease and body habitus (standardised beta 0.315, p=0.012). Conversely, PORH did not correlate with CMR perfusion at rest or after stress.

**Conclusions:** Maximal hyperaemic response is associated with resting CMR perfusion independent of conventional risk factors. This suggests that cardiac microvascular dysfunction may represent a manifestation of wider microcirculatory derangements. Further research is required to determine whether interventions that improve systemic vascular disturbances may improve cardiac microcirculation.

**Translational Perspective:** It is recognised that coronary microvascular dysfunction is associated with residual symptoms in people with angina, after the correction of occlusive coronary arterial disease. As such it is a promising target for symptom control, however development of proof-of-concept trials is limited by the ability to monitor the coronary microcirculation in those trials.

This manuscript identifies an appropriate surrogate endpoint that can be easily and non-invasively monitored and validates it against MRI imaging of the coronary microcirculation.

## Introduction

Coronary microvascular dysfunction (CMD) is usually defined in the context of angina with non-obstructed coronary arteries (ANOCA)^6, 17, 31^, although it is increasingly recognised that it co-exists with epicardial coronary artery disease^7, 8, 12^. This belies the fact that CMD carries a negative prognostic impact, independent of the presence of focal coronary artery stenosis^41^. Accordingly, the diagnosis of CMD is important in guiding prognostication over and above coronary angiographic appearance.

Coronary microvasculature structure and function can be quantified either invasively or non-invasively. Whilst evaluation of the role of CT coronary angiography (CTCA) in the assessment and treatment of CMD is underway, the gold-standard remains invasive assessment^36^. Invasive angiographic methods can be used to characterise coronary microvascular function^19, 29, 32^ despite the burgeoning demand for non-invasive chest pain investigation pathways^15^ and the evidence for those pathways’ safety and efficacy^13, 22^. This invasive testing is not without risk to the patient, and there is a need to establish non-invasive methods of investigation.

The non-invasive options for investigating CMD are usually selected from one or more of: positron emission tomography (PET), Doppler echocardiography, or cardiovascular magnetic resonance imaging (CMR)^40^. Whilst all these techniques have relative advantages and deficiencies, the prevalence of CMR investigation in current clinical practice, combined with its flexibility (providing tissue, structural, and perfusion/functional imaging), make it an attractive option for further study^4^.

Skin microcirculation is an established model to investigate systemic microvascular function prior to the clinical manifestation of disease^26^. It has been independently associated with symptomatic chest syndromes^38^, renal disease^37^, retinopathy^30^, and some measures of cardiac function^18, 26^. However, the exact relationship between peripheral and cardiac measures of microcirculatory function is complicated by the wide array of techniques on both sides, and some conflicting results^27^. Additionally, the cohorts examined have almost universally concentrated on patients with angina with non-occlusive coronary artery disease (ANOCA: usually defined as <50% stenosis of the coronary arteries ^4, 16, 19, 32^).

We aimed to examine the relationship between peripheral and coronary microvascular function using laser Doppler fluximetry and CMR respectively. We aimed to do this in populations both with and without coronary artery disease, as assessed using CTCA.

## Methods

### Study population

This study conformed to the Declaration of Helsinki and was approved by the National Research Ethics Service Southwest (10/H0206/67 and 10/H0102/66). All participants gave informed written consent.

Participants were prospectively recruited from the cardiology services at two centres, the Royal Devon and Exeter Hospital (part of the Royal Devon and Exeter NHS Foundation trust, Exeter, UK) and Derriford Hospital, (part of the University Hospitals Plymouth NHS Trust, Plymouth UK), and from a population-based sample enriched for people with type 2 diabetes mellitus. Those recruited from the cardiology services could have either proven atherosclerotic heart disease or diagnosed microvascular angina (non-obstructive coronary arteries assessed via CTCA). Coronary artery disease (CAD) was defined as a coronary artery calcium score on CTCA greater than 0 as suggested by contemporary research and guidelines on lipid management.^5, 21, 25^ This included a population who self-reported previous diagnosis of atherosclerosis, coronary revascularisation or acute coronary syndromes, and a further subgroup who had no clinical manifestations of coronary artery disease, but CTCA demonstrated significant atherosclerosis. After written informed consent was obtained, basic demographics, including medication history and biochemical and haematological measures of cardiovascular risk were recorded. Subjects underwent cardiac imaging with CTCA, CMR and systemic microvascular assessments.

### Cardiac imaging

#### CTCA

CTCA was performed using a multidetector scanner at the Royal Devon & Exeter or Derriford Hospital and reported blind to location or clinical characteristics by two experienced clinicians. Stenoses were visually assessed and graded using a five-point severity scale (0 to 4 corresponding to no atheroma, <20%, 20-50%, 51-70%, and >70% respectively). Coronary artery calcium Agatston scoring was performed using standard protocols^3^.

#### CMR

A 1.5T Philips MRI scanner was used. Baseline acquisition scans were obtained. Intravenous gadolinium-DTPA contrast agent was delivered and flushed with saline via an intravenous cannula. Three short axis slices (basal, mid-ventricular and apical) were acquired using a gradient-echo pulse sequence and regional myocardial perfusion assessment was performed according to the American Heart Association 16-segment model. A continuous intravenous infusion of adenosine (140mcg/min/kg for 4-6 minutes via a second intravenous cannula) was administered and imaging was repeated. Regional qualitative, semi-quantitative and absolute myocardial perfusion estimates were derived at baseline and maximal vasodilatory stimulus. A myocardial perfusion index (MPI) at stress (MPIstress) and at rest (MPIrest) was calculated using the ratios of the gradients of signal intensity over time in myocardial segments to left ventricular blood pool. Myocardial perfusion reserve index (MPRI) was calculated using the ratio of MPIstress to MPIrest:

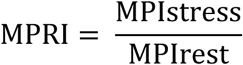

both for subendocardial (inner) and subepicardial (outer) halves of segments.

### Peripheral microvasculature

As skin perfusion can be highly variable at rest, microvascular function was assessed by its response to fixed stimuli, namely maximal hyperaemic response (MHR) to local heating and post occlusive reactive hyperaemia (PORH).

#### Skin maximal hyperaemic response

Methods were used as previously described by Adingupu D et al.^1^. A small brass heater (0.76cm^2^) (Moor Instruments, Axminster UK) was attached to the dorsal surface of the foot and heated the skin to 42°C for 30 minutes. Skin perfusion was then assessed in 8 equidistant sites by laser Doppler fluximetry of (DRT4 system, Moor Instruments, Axminster, Devon, U.K.). The mean of these eight sites represents MHR. Minimum vascular resistance (MVR) was calculated via the equation:

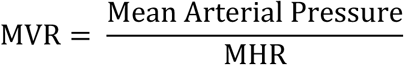

#### Post-occlusive reactive hyperaemia

A multi-outlet blood pressure cuff, connected to a large air reservoir was placed around the ankle according to a standardised protocol^1^. A laser Doppler probe (780nM) (DRT4 system, Moor Instruments, Axminster, Devon, U.K.) attached to the dorsal surface of the foot was used to take baseline recordings for 4 minutes in the supine position. The cuff was then inflated to a minimum of 200mmHg or 20mmHg higher than systolic blood pressure for 4 minutes following which it was rapidly deflated, and blood flow recorded for 10 minutes post release. Peak response and time to peak response were evaluated with a time constant of 0.1 second.

### Statistical analysis

Variables of interest were assessed for normality using Kolmogorov-Smirnov testing and, when possible, appropriately transformed to allow parametric analysis. Mean and standard deviations (SD) were calculated and compared using two-sided Student’s t-test. Comparisons according to presence or absence of both coronary artery disease (as defined above) and diabetes were made. Geometric means and interquartile ranges calculated are presented for transformed data. Categorical variables were compared using Pearson’s Chi-Squared testing.

To explore predictors of MRI derived perfusion indices, multivariable linear regression analyses were performed. All potential variables with an *a priori* defined hypothesis were tested in a prioritised order as determinants. Standardised beta coefficients (change in parameter per SD change of the variable) are presented alongside p-values, with p<0.05 considered statistically significant. Statistical analyses were performed using Stata v17.0 (StataCorp, College Station, TX; USA).

## Results

102 subjects underwent the microvascular assessments, CMR, and CTCA. Patient demographic characteristics, CAC scores and biochemical markers of cardiovascular risk results are summarised in table 1.

**Table 1.** Baseline characteristics (mean (standard deviation) or geometric mean [25th – 75th quartiles]) of the cohort stratified by the presence of Coronary artery disease (CAD) defined as coronary artery calcification score >0 Agatson units or previous history of MI or myocardial revascularisation procedure.

| *Variable* | *All pts (n=102)* | *CAD (n=46)* | *No CAD (n=56)* | *p value* |
| --- | --- | --- | --- | --- |
| *Age, (years)* | 63.2 (9.4) | 68.0 (7.2) | 59.3 (9.3) | 0.0004 |
| *Sex (% male)* | 60.8% | 59.7% | 40.3% | 0.0008 |
| *Weight (kg)* | 81.9 (16.5) | 85.1 (15.7) | 79.3 (16.8) | 0.07 |
| *BMI (kg/m<sup>2</sup>)* | 28.3 (4.7) | 28.5 (4.2) | 28.0 (5.0) | 0.6 |
| *Systolic BP (mmHg)* | 138.4(14.4) | 140.0 (14.2) | 137.4 (14.6) | 0.4 |
| *Diastolic BP, (mmHg)* | 80.1 (8.9) | 78.7 (9.4) | 81.3 (8.3) | 0.1 |
| *Total Cholesterol (mmol/l)* | 4.7 (3.8-6.1) | 4.2 (3.5-4.9) | 5.1 (4.4 – 6.2) | 0.001 |
| *HDL cholesterol (mmol/l)* | 1.5 (1.2-1.9) | 1.3 (1.1-1.5) | 1.6 (1.3-2.0) | 0.004 |
| *LDL cholesterol (mmol/l)* | 2.5 (1.9-3.7) | 2.1 (1.7-2.8) | 2.8 (2.3-3.9) | 0.004 |
| *Triglycerides (mmol/l)* | 1.2 (0.9-1.5) | 1.2 (0.9-1.6) | 1.2 (0.9-1.5) | 0.8 |
| *HbA1c (mmol/mol)* | 45 (38-54) | 49 (40-58) | 43 (37-48) | 0.002 |
| *eGFR, (mls/min)* | 80 (72-90) | 79 (74-90) | 81 (71-90) | 0.4 |
| *Type 2 Diabetes (%)* | 45.1% | 52.5% | 34.9% | 0.08 |
| *Previous MI (%)* | 2.9% | 5.1% | 0% | 0.1 |
| *Previous cerebrovascular event (%)* | 5.0% | 6.8% | 2.4% | 0.3 |
| *Smoker (%)* | 52.5% | 50.8% | 54.8% | 0.7 |
| ***Medications.*** |  |  |  |  |
| *Betablocker* | 9.8% | 11.9% | 7.0% | 0.4 |
| *CCB* | 15.7% | 18.6% | 11.6% | 0.3 |
| *ACEi* | 21.6% | 32.2% | 7.0% | 0.002 |
| *Statin* | 54.9% | 64.4% | 41.9% | 0.02 |
| *Antiplatelet* | 28.4% | 35.6% | 18.6% | 0.06 |
| *CAC score, (AU)* | 17 (0-292) | 326 (118-807) | 0.0 (0.0-0.0) | <0.0001 |

MI, myocardial infarction; BMI, body mass index; CCB, calcium channel blocker; ACE-I, angiotensin converting enzyme inhibitor; CAC, coronary artery calcium. Measured in agatston units (AU).

Those with CAD were older, were more likely to be male and have a prior diagnosis of hypertension. Those with CAD were also more likely to be on diabetic or statin treatment or have an ACE inhibitor prescribed. There was no difference in blood pressure or serum lipids according to CAD status, though this may be explained by more statin and antihypertensive use in the CAD group.

### Skin microvascular assessment

In keeping with previous reports, participants with diabetes had attenuated MHR and an increased MVR compared to those without diabetes (mean MHR 183 (66.2) vs. 229 (63.7) arbitrary units (au) for those with and without diabetes respectively, p<0.001; table 2). There were no statistically significant differences amongst any of the parameters measuring peripheral microvascular function when grouped according to presence or absence of coronary artery disease (table 2).

**Table 2.**
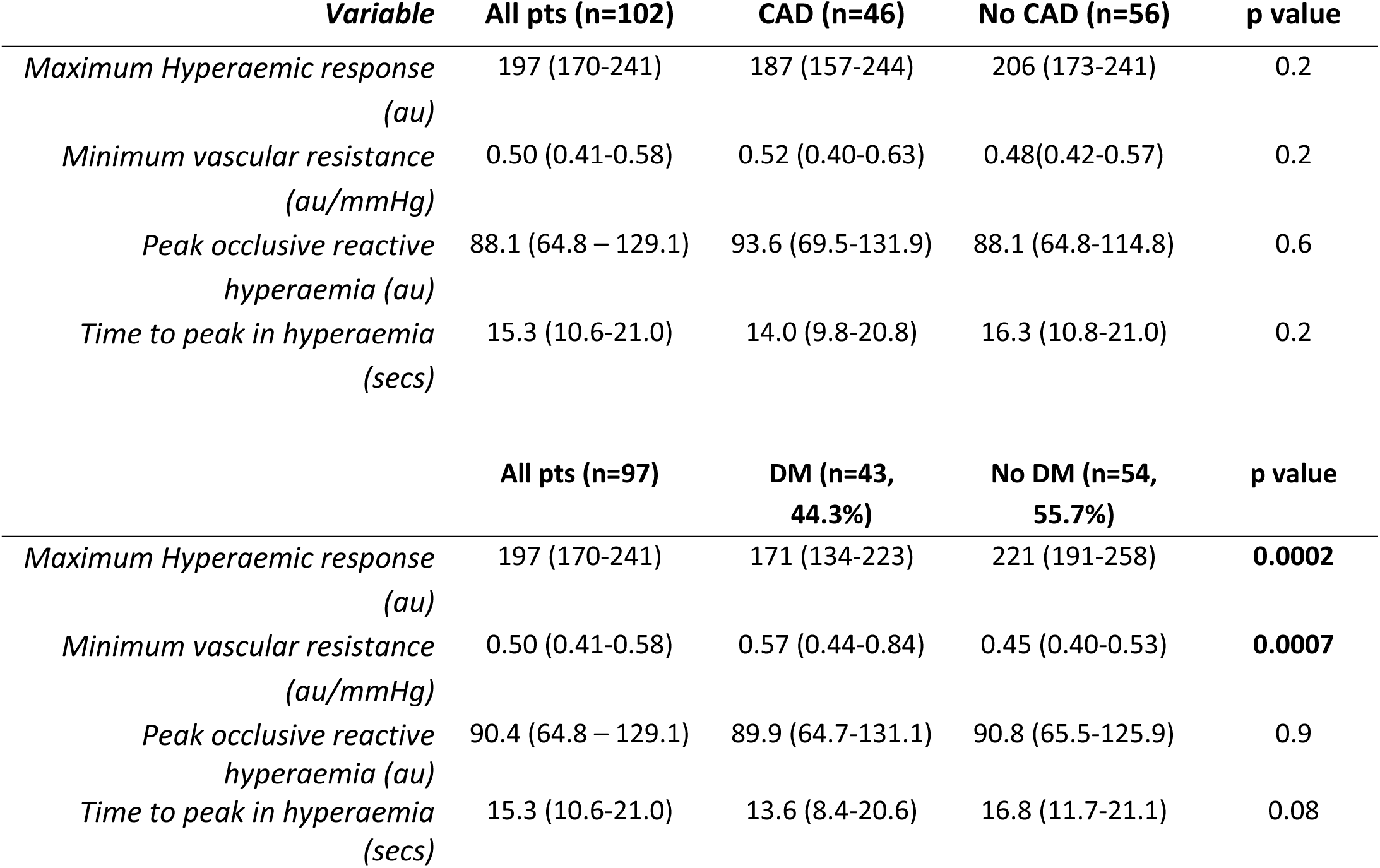
Peripheral microvascular measurements analysed grouped according to coronary artery and diabetic disease status. Data are presented as geometric means with interquartile ranges.

| **Variable** | **All pts (n=102)** | **CAD (n=46)** | **No CAD (n=56)** | **p value** |
| --- | --- | --- | --- | --- |
| *Maximum Hyperaemic response (au)* | 197 (170-241) | 187 (157-244) | 206 (173-241) | 0.2 |
| *Minimum vascular resistance (au/mmHg)* | 0.50 (0.41-0.58) | 0.52 (0.40-0.63) | 0.48(0.42-0.57) | 0.2 |
| *Peak occlusive reactive hyperaemia (au)* | 88.1 (64.8 – 129.1) | 93.6 (69.5-131.9) | 88.1 (64.8-114.8) | 0.6 |
| *Time to peak in hyperaemia (secs)* | 15.3 (10.6-21.0) | 14.0 (9.8-20.8) | 16.3 (10.8-21.0) | 0.2 |
|  | **All pts (n=97)** | **DM (n=43, 44.3%)** | **No DM (n=54, 55.7%)** | **p value** |
| *Maximum Hyperaemic response (au)* | 197 (170-241) | 171 (134-223) | 221 (191-258) | **0.0002** |
| *Minimum vascular resistance (au/mmHg)* | 0.50 (0.41-0.58) | 0.57 (0.44-0.84) | 0.45 (0.40-0.53) | **0.0007** |
| *Peak occlusive reactive hyperaemia (au)* | 90.4 (64.8 – 129.1) | 89.9 (64.7-131.1) | 90.8 (65.5-125.9) | 0.9 |
| *Time to peak in hyperaemia (secs)* | 15.3 (10.6-21.0) | 13.6 (8.4-20.6) | 16.8 (11.7-21.1) | 0.08 |

### Cardiac MRI results

There were no significant differences in MRI myocardial perfusion parameters, either at rest or at stress, in those with diabetes. Resting myocardial perfusion was reduced in those with coronary artery disease compared to those without (table 3).

**Table 3.** Cardiac MRI perfusion measurements analysed grouped according to coronary artery and diabetic disease status after adjustment for age and sex. Data are presented as geometric means with 95% Confidence intervals

| *Variable* | *All pts (n=75)* | *CAD (n=42, 56%)* | *No CAD (n=33, 44%)* | *p value* | *DM (n=30, 37%)* | *No DM (n=47, 63%)* | *p value* |
| --- | --- | --- | --- | --- | --- | --- | --- |
| ***MPI under stress*** |  |  |  |  |  |  |  |
| *Subendocardial* | 0.130 (0.121-0.140) | 0.118 (0.104-0.140) | 0.140 (0.124-0.158) | 0.1 | 0.127 (0.112-0.145) | 0.132 (0.119-0.146) | 0.7 |
| *Subepicardial* | 0.138 (0.128-0.150) | 0.127 (0.111-0.146) | 0.148 (0.131-0.167) | 0.2 | 0.134 (0.117-0.154) | 0.141 (0.127-0.156) | 0.6 |
| *Whole segment* | 0.139 (0.129-0.150) | 0.126 (0.106-0.149) | 0.151 (0.135-0.169) | 0.06 | 0.137 (0.121-0.156) | 0.141 (0.127-0.155) | 0.8 |
| ***MPI at rest*** |  |  |  |  |  |  |  |
| *Subendocardial* | 0.084 (0.077-0.091) | 0.070 (0.061-0.081) | 0.097 (0.085-0.110) | 0.005 | 0.074 (0.064-0.086) | 0.080 (0.071-0.089) | 0.5 |
| *Subepicardial* | 0.078 (0.071-0.084) | 0.068 (0.059-0.079) | 0.086 (0.075-0.099) | 0.04 | 0.075 (0.064-0.090) | 0.079 (0.057-0.108) | 0.6 |
| *Whole segment* | 0.082 (0.075-0.089) | 0.070 (0.061-0.082) | 0.092 (0.081-0.105) | 0.02 | 0.079 (0.068-0.091) | 0.083 (0.074-0.094) | 0.5 |
| ***MPRI*** |  |  |  |  |  |  |  |
| *Subendocardial* | 1.631 (1.493-2.072) | 1.714 (1.477-1.990) | 1.565 (1.370-1.789) | 0.4 | 1.632 (1.411-1.888) | 1.631 (1.450-1.834) | 0.99 |
| *Subepicardial* | 1.880 (1.718-2.059) | 1.917 (1.646-2.232) | 1.851 (1.615-2.122) | 0.8 | 1.850 (1.596-2.146) | 1.902 (1.688-2.142) | 0.8 |
| *Whole segment* | 1.748 (1.603-1.906) | 1.803 (1.560-2.083) | 1.704 (1.495-1.942) | 0.6 | 1.764 (1.532-2.032) | 1.738 (1.550-1.949) | 0.9 |

CAD = coronary artery disease (defined as per table 1); MPI = myocardial perfusion index; MPRI = myocardial perfusion reserve index = MPI<sub>stress</sub>/MPI<sub>rest</sub>.

There was an association between maximum hyperaemic response, minimum microvascular resistance and resting myocardial perfusion such that for every standard deviation increase in skin microvascular response, the myocardial perfusion increased by approximately a third (figure 1). These associations were present to similar degrees in sub-endocardial, sub-epicardial and the full thickness of the myocardium. Multivariate regression for potential confounders and mechanistic associations did little to reduce these associations (table 4). There was no association between MPI under stress and peripheral microvascular perfusion.

**Figure 1.**
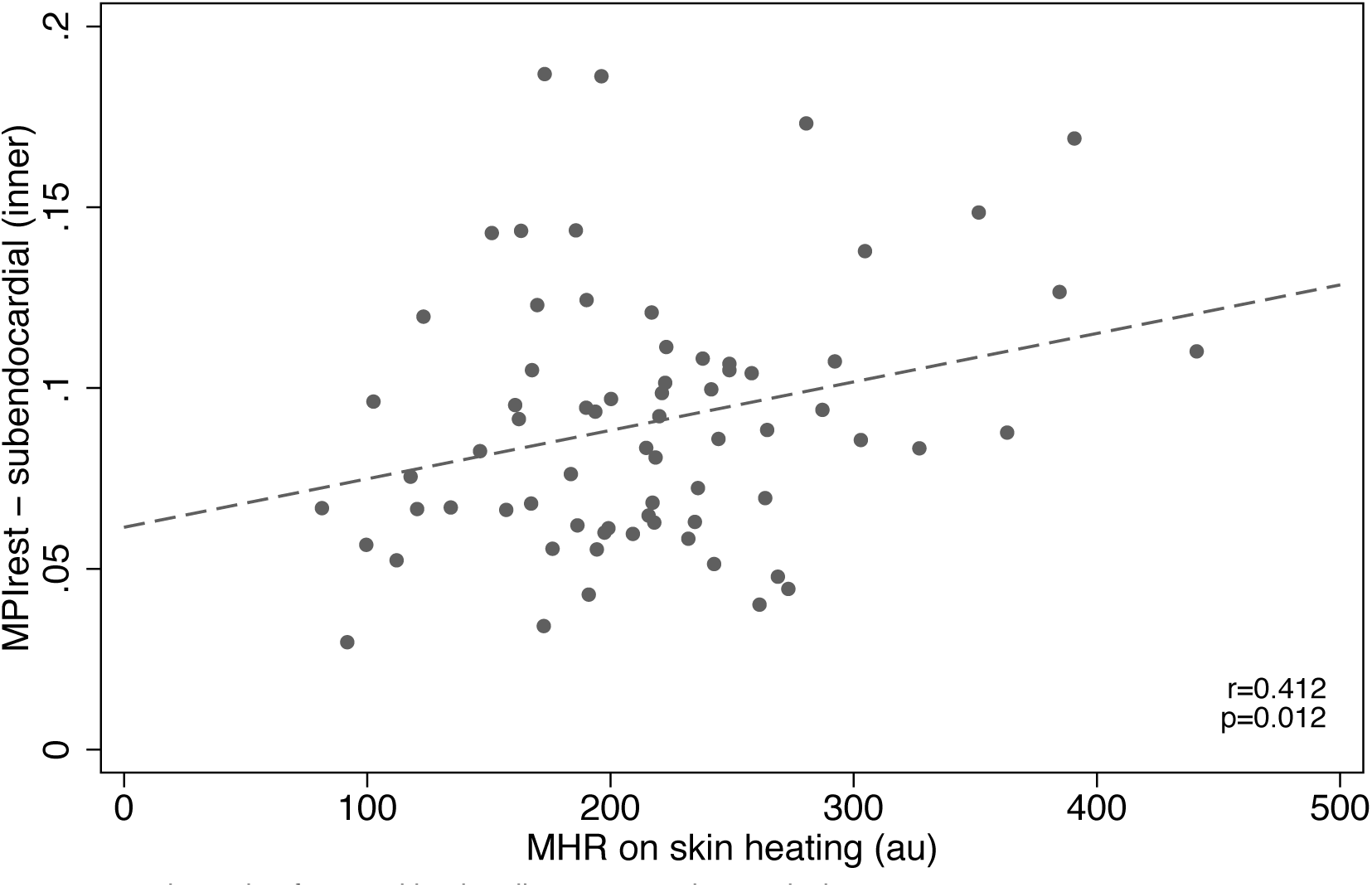
Association between subendocardial myocardial perfusion index with the maximum hyperaemic response in the skin after adjustment for age, sex, presence of diabetes and systolic blood pressure.

**Table 4.**
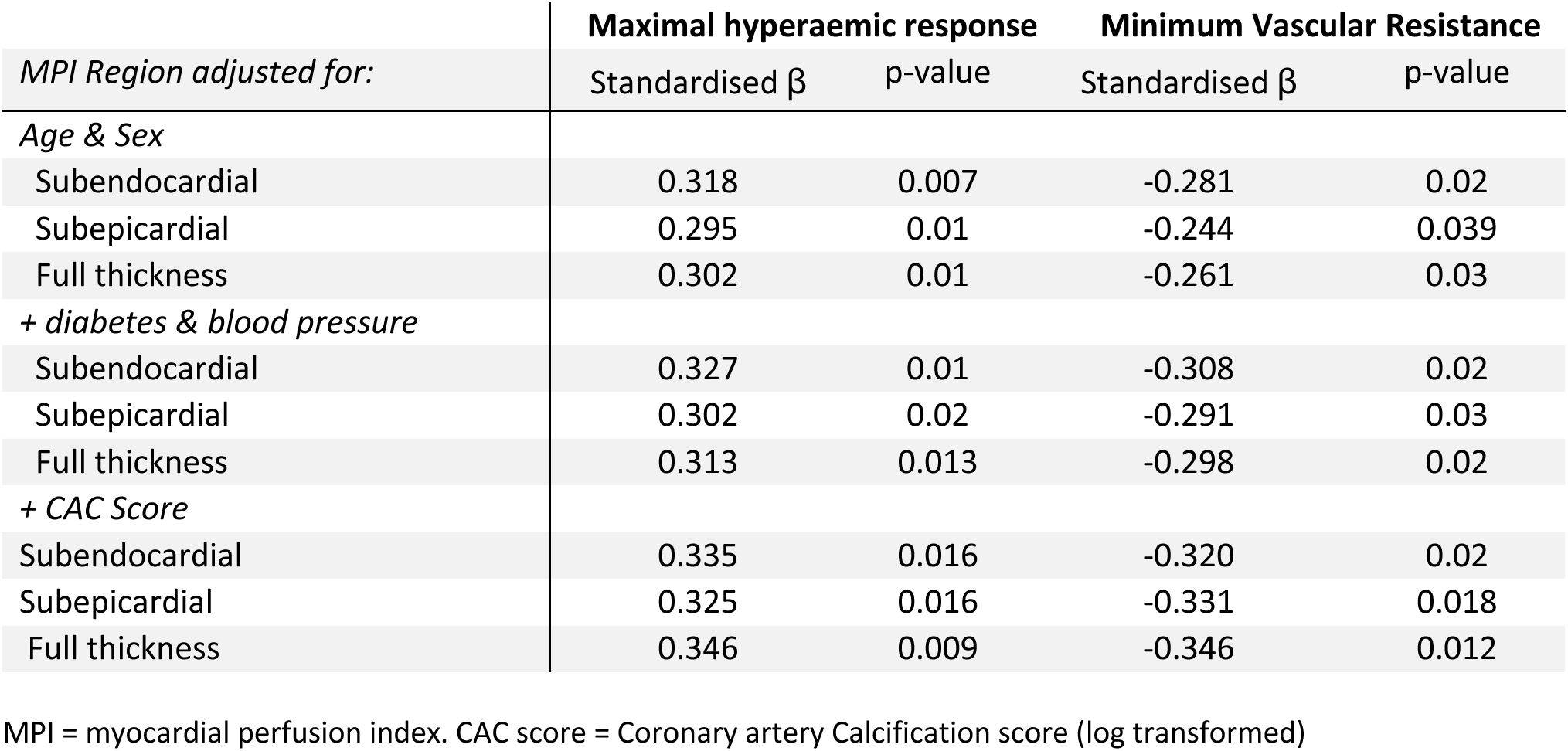
Associations between peripheral microvascular parameters (maximum hyperaemic response and minimum microvascular resistance) and resting cardiac microvascular perfusion. Results are presented as standardised-betas (i.e. change in parameter per standard deviation change in the variable) with a negative value representing an inverse correlation

| *MPI Region adjusted for:* | **Maximal hyperaemic response** |  | **Minimum Vascular Resistance** |  |
| --- | --- | --- | --- | --- |
| | Standardised $\beta$ | p-value | Standardised $\beta$ | p-value |
| *Age & Sex* |  |  |  |  |
| Subendocardial | 0.318 | 0.007 | -0.281 | 0.02 |
| Subepicardial | 0.295 | 0.01 | -0.244 | 0.039 |
| Full thickness | 0.302 | 0.01 | -0.261 | 0.03 |
| *+ diabetes & blood pressure* |  |  |  |  |
| Subendocardial | 0.327 | 0.01 | -0.308 | 0.02 |
| Subepicardial | 0.302 | 0.02 | -0.291 | 0.03 |
| Full thickness | 0.313 | 0.013 | -0.298 | 0.02 |
| *+ CAC Score* |  |  |  |  |
| Subendocardial | 0.335 | 0.016 | -0.320 | 0.02 |
| Subepicardial | 0.325 | 0.016 | -0.331 | 0.018 |
| Full thickness | 0.346 | 0.009 | -0.346 | 0.012 |

MPI = myocardial perfusion index. CAC score = Coronary artery Calcification score (log transformed)

When considering the myocardial perfusion reserve index (MPRI) there was a paradoxical inverse correlation such that for an increased peripheral response there was an attenuation of MPRI (Standardised beta -0.271; p=0.025 for correlation between MHR and subendocardial MPRI), however this was driven by the positive association between skin perfusion and resting myocardial perfusion and an absence of association with stress response. Adjustment for coronary artery calcification did not substantially alter the magnitude nor the significance of the association between systemic microvascular response and MPI at rest in any cardiac territory.

## Discussion

We have demonstrated or the first time that resting coronary microvascular perfusion correlates with systemic circulation, such that lower skin microvascular perfusion was very closely associated with impaired resting coronary perfusion. These results represent the only published data to our knowledge comparing human coronary microvascular structure and function using cardiac MRI and CTCA with peripheral microvascular structure and function assessed by laser Doppler fluximetry. This association between systemic and coronary disease is particularly pertinent given the current emphasis on non-invasive delineation of coronary anatomy with both outcomes^22, 45^ and guidelines^15^.

The current focus on systemic disease is based on previous cross-sectional work that implied, but did not actually demonstrate, that systemic disease may also impact the coronary microcirculation. The literature has reported abnormalities in the systemic microcirculation in cross sectional studies comparing patients with coronary disease and healthy controls, as in this comparison of patients with early angina, obstructive coronary artery disease or acute coronary syndromes with healthy controls^31^.

Ford et al. described differences in biopsied gluteal resistance artery function amongst groups of patients with invasively diagnosed coronary microvascular angina (MVA) and chest pain with neither obstructive coronary artery disease nor MVA as controls.^18^. This study, though in vessels larger than the microcirculation, suggested that systemic abnormalities of small blood vessels may be apparent in those with microvascular angina, in particular impaired relaxation and enhanced vasoconstrictor responses to vasoactive agents^39^. Future CorMicA sub-studies hold the potential to relate peripheral vascular resistance artery function to CMR and CTCA^11, 36^, but these will not directly study the microcirculation as was undertaken in our study.

In healthy volunteers, Khan et al. showed that peripheral microvascular function measured by laser Doppler fluximetry after iontophoretic application of acetylcholine and sodium nitroprusside was positively correlated with coronary flow reserve measured by transthoracic echocardiography with intravenous adenosine^26^, indicating that in health the skin microcirculation is representative of more generalised microvascular function including the coronary microcirculation.

Shamim-Uzzamam et al. showed that brachial artery flow-mediated dilatation is unrelated to PORH in healthy controls and CAD patients, but that time to peak response in PORH differed amongst CAD patients versus controls^35^. Exploring this in our data, a sensitivity analysis using increasing CAC score cut-offs for our definition of CAD showed that MVR showed significant differences between CAD and no CAD groups (CAC score >60, t-test p<0.05), but given the increasing acceptance of CAC score 0 to indicate no CAD in the literature^5^ and guidelines^21^ we felt an arbitrary cut off based on CAD>60 would be less transferable to other studies/clinical practice. Clearly the presence of people with diabetes in both our CAD positive and negative cohort also makes our groups not directly comparable to those of Shamim-Uzzamam.

Agarwal et al. studied PORH, MHR, and iontophoresis amongst patients with angiographically proven CAD, finding impaired responses compared to healthy age-and sex-matched controls^2^. However, CAD patients were likely recruited after a diagnosis of symptomatic cardiovascular disease: this, combined with the health of the controls, explains the magnitude of the difference between groups compared to our study.

Van der Heijden et al. examined PORH in patients undergoing coronary angiography^43^, finding BMI correlated with impaired cutaneous microvascular function as measured by laser Doppler fluximetry, peripheral arterial tonometry, and digital thermal monitoring . Our data partially support this finding in both univariate (p=0.007, standardised beta=-0.273) and multivariate (p=0.050, standardised beta=-0.213) regression analysis of MHR with BMI (and age, sex, diabetes, CAC score and systolic BP as covariates), though neither PORH nor any MRI parameter was found to be correlated to BMI. Differences between measurement definition and technique may account for the discordant PORH results, though the agreement on maximum hyperaemia adds external validity to our data.

We have demonstrated a positive correlation between skin microvascular MHR and resting cardiac MRI parameters of perfusion. These associations suggest that a lower maximium vasodilatory capacity of the skin microcirculation is available in those subjects with lower resting levels of myocardial perfusion. Resting cardiac perfusion defects have been associated with cardiac transplant arteriopathy^28^, and may predict poor cardiovascular outcomes: a non-invasive peripheral microvascular surrogate of resting cardiac perfusion may have clinical utility in the diagnosis and monitoring of cardiovascular disease, however further research is needed to explore this hypothesis.

Heart failure is a chronic progressive disease, with no known cure, which is characterised by the heart’s inability to meet the perfusion requirements of the body. Although early studies suggested this was due to atherosclerotic disease, revascularisation strategies provided little additional benefit over optimal medical therapy in the STICH and HEART trials^10, 44^ and by experimental models^20^. There is now a growing recognition that cardiac microvascular impairment is at the root of much heart failure^14, 34, 42^. As the myocardium spends the majority of time in a resting state, the demonstrated correlation between peripheral microvascular disease and resting cardiac perfusion on MRI provides further evidence that this is a systemic rather than a local problem. This is an important consideration when exploring preventative strategies. We have previously shown that skin microvascular pathology responds over time to improvements in baseline parameters such as body weight^9^. This provides circumstantial evidence that the benefit that others have demonstrated in weight loss reducing incident heart failure may be, in part, mediated through global microcirculatory function improvement. This will require further investigation in longitudinal studies.

We found that MHR was statistically significantly lower for participants with than those without diabetes as previously reported ^23, 24, 46^, however no MRI parameter was found to be different between these groups. Despite thorough exploratory analyses we were unable to establish any confounding variables to explain this. It is important to clarify that those living with diabetes in this cohort were “healthier” than the typical population, they had excellent control (median HbA1c 55.5mmol/mol, IQR 47-62mmol/mol) and a relatively short duration of diagnosis. These patients also have minimal clinical microvascular complications, with essentially normal eGFR, a low prevalence of microalbuminuria and retinopathy, and their peripheral microvascular functional abnormalities are limited. Further research should explore whether cardiac microvascular abnormalities are evident in diabetics with more extensive peripheral microcirculatory dysfunction and/or clinically evident microvascular complications.

The lack of differentiation between participants with and without CAD amongst their stress perfusion MRI results is likely explained by the degree and proportion of epicardial coronary stenosis amongst this cohort. Only 14 (13%) of our participants had any stenosis greater than 70%, compared with 48.8% of participants with stenoses ≥ 75% in MR-IMPACT II^33^. Similarly, the prevalence of diagnosed cardiovascular disease was low in our cohort, with only 6 out of 106 participants reporting a diagnosis of PCI, CABG, acute coronary syndrome or other cardiovascular disease. Further studies in populations enriched for coronary artery disease risk factors may reveal stronger relationships – particularly where angiography reveals no coronary stenosis – between cardiac and peripheral microvascular parameters.

## Limitations

This is an observational study without details of clinical outcomes, and is hypothesis generating rather than predictive. Additionally, 102 participants represent a small cohort, especially considering only 45% were diabetic and 3% had had a previous acute coronary syndrome.

Invasive coronary angiography would have given greater certainty around stenosis numbers and severity. However the heavy emphasis of CTCA in contemporary guidelines^15^ means our cohort aligns well with clinical practice, and allows direct applicability of any future outcome studies.

## Conclusion

The maximal hyperaemic response (MHR) and minimum vascular resistance are non-invasive peripheral measures of skin microcirculation and correlate with resting measures of perfusion on cardiac MRI. Other alternative measures of peripheral microvascular function, namely post-occlusive reactive hyperaemia, were found to be unrelated to CMR perfusion. The correlation between resting cardiac perfusion measures persisted even after multivariate regression analyses including CAC score.

## Data Availability

All data produced in the present study are available upon reasonable request to the authors.

## Declarations

### Consent for publication

Not applicable

### Availability of data and materials

Fully anonymised data are available by direct application to the corresponding author.

### Competing interests

The authors declare that they have no competing interests.

### Funding

This work was supported by the Innovative Medicines Initiative (the SUMMIT consortium, IMI-2008/115006). OG was funded by the Gawthorn Cardiac Trust.

### Authors Contributions

ACS, KMG, OG, NB, and WDS were involved in study design and securing funding. <u>KMG, FC</u> <u>OG</u> were involved in study management and data collection. ACS, NB and DS supervised the project delivery. JW, JH, FC, OG and WDS contributed to database analysis. JW, ACS and WDS performed the statistical analysis and produced the primary manuscript. All authors have seen and contributed to the drafting and revision of the manuscript and have approved the final version for publication. WDS is responsible for the integrity of the work as a whole.

## Acknowledgements

This work was supported by the Innovative Medicines Initiative (the SUMMIT consortium, IMI-2008/115006). OG was funded by The Gawthorn Cardiac Trust fellowship. JW was supported by an academic clinical fellowship with the NIHR. This paper presents independent research supported by the NIHR Exeter Clinical Research Facility and the NIHR Collaboration for Leadership in Applied Health Research and Care (CLAHRC) for the South West Peninsula. The views expressed in this publication are those of the author(s) and not necessarily those of the NIHR Exeter Clinical Research Facility, the NHS, the NIHR or the Department of Health and social care in England. We would also like to acknowledge and thank the South West Stroke Research Network for their help with patient recruitment, and all of the patients who took part in this research.

## Notes

### Competing Interest Statement

The authors have declared no competing interest.

